# A wake-up call - revealing the oversight of sleep physiology and related translational discrepancies in studies of rapid-acting antidepressants

**DOI:** 10.1101/2020.09.29.20204008

**Authors:** Okko Alitalo, Roosa Saarreharju, Carlos A. Zarate, Samuel Kohtala, Tomi Rantamäki

## Abstract

Depression and sleep problems go hand-in-hand, while clinical improvement often emerges along the normalization of sleep architecture and realignment of the circadian rhythm. Antidepressant effects of sleep deprivation and cognitive behavioral therapy targeted at insomnia further demonstrate the confluence of sleep and mood. Moreover, recent literature showing that ketamine influences many processes related to sleep-wake neurobiology, have led to novel hypotheses explaining rapid and sustained antidepressant effects. Surprisingly, studies addressing ketamine’s antidepressant effects have had a narrow focus on solely on pharmacological aspects and often ignore the role of physiology. To illustrate this discrepancy, we conducted a literature review on articles around rapid-acting antidepressants published between 2009-2019. A gross keyword check indicated overall ignorance of sleep in most studies. To investigate the topic closer, we focused on the most cited preclinical and clinical research papers. Circadian rhythm, timing of drug administration and behavioral tests relative to light cycles, sleep, and their potential association with experimental observations were mentioned only in a handful of the papers. Most importantly, in preclinical reports the treatments have been preferentially delivered during the inactive period, which is polar opposite to clinical practice and research. We hope this report serves as a wake-up call for sleep in the field and urges (re)examining rapid-acting antidepressant effects from the perspective of wake-sleep physiology.

## 1. Introduction

The glutamatergic *N*-methyl-D-aspartate receptor (NMDAR) antagonist ketamine is a dissociative anesthetic drug with rapid-acting antidepressant properties. The discovery of ketamine’s antidepressant effects has been described as one of the most important advances in modern psychiatry, since traditional antidepressants often require weeks or months of use before depressive symptoms are efficiently relieved. A single dose of ketamine often results in rapid (within hours) reduction of depressive symptoms in individuals suffering from major depressive disorder (MDD) (Berman et al., 2000), bipolar depression (Diazgranados et al., 2010; Zarate et al., 2012), and treatment-resistant depression (Fava et al., 2020; Murrough et al., 2013; Zarate et al., 2006). Recent studies indicate that repeated dose intravenous ketamine may significantly prolong antidepressant effects (Park et al., 2019). While ketamine is generally well tolerated, it is a drug of abuse with a rich profile of psychotomimetic effects that complicate its widespread use (Horowitz & Moncrieff, 2020). Moreover, a subset of individuals do not achieve remission despite repeat dosing of ketamine (Phillips et al., 2019). These limitations have prompted investigation into finding possibilities for extending and reinforcing ketamine’s therapeutic effects and discovering more effective and longer lasting alternatives.

Extensive research efforts have been invested to uncover the mechanisms governing ketamine’s antidepressant effects (for review see: Rantamäki & Kohtala, 2020; Zanos, et al., 2018). Recent studies have pinpointed the effects of ketamine on the increased release of the excitatory neurotransmitter glutamate within the medial prefrontal cortex and some subcortical structures, activation of α-amino-3-hydroxy-5-methyl-4-isoxazolepropionic acid receptors (AMPARs) and subsequent facilitation of BDNF (brain-derived neurotrophic factor) dependent synaptic plasticity and synaptogenesis (Deyama & Duman, 2020; Duman et al., 2016; Duman & Aghajanian, 2012; Rantamäki & Yalcin, 2016). An alternative, or complementary, proposed mechanism by which ketamine increases BDNF signaling and synaptic plasticity transpires from the blockade of synaptic NMDARs involved in spontaneous synaptic transmission, which deactivates eukaryotic elongation factor 2 kinase (eEF2K), resulting in dephosphorylation of eEF2 and the subsequent de-suppression of BDNF protein synthesis (Monteggia et al., 2013). Notably, other treatments associated with rapid antidepressant effects, such as electroconvulsive therapy (ECT) and sleep deprivation, also produce cellular and molecular changes analogous to ketamine (Rantamäki & Kohtala, 2020). Despite major advances in understanding the molecular basis underlying ketamine’s effects, the past two decades of drug discovery have not been able to produce compounds with matching therapeutic effects. While several successful rapid-acting drug candidates have demonstrated efficacy in preclinical setting, they have subsequently failed in clinical trials (Garay et al., 2018; Kadriu et al., 2019). The lack of success for antidepressant drugs has been traditionally explained with the poor construct validity of animal models of depression (Cryan & Holmes, 2005; Harmer et al., 2011; Hasler et al., 2004; Nestler et al., 2002). However, it has become increasingly evident in translational brain research that many other features, such as selection of animal species and strains, age of the animals, sex, and even microbiota, can have a major impact on study outcomes and should be properly accounted and controlled in experimental design (Cryan & Holmes, 2005; Franklin & Ericsson, 2017; Võikar et al., 2001). But have we perhaps forgotten something even more fundamental to animal physiology?

Depressive disorders are inherently associated with disrupted sleep and circadian rhythms (Hasler et al., 2004; Nutt et al., 2008). On the other hand, treatments like sleep deprivation produce rapid, although often highly transient, antidepressant effects. Most importantly, recent studies have associated homeostatic and circadian components of sleep with the antidepressant effects of ketamine (Duncan et al., 2013, 2017, 2018; Kohtala et al., 2019). These and other studies have sparked several notable hypotheses with the goal of explaining the neurobiological and behavioral outcomes of rapid-acting antidepressant treatments by inspecting them in the context of sleep and/or circadian rhythm (Duncan et al., 2019; Orozco-Solis et al., 2017; Rantamäki & Kohtala, 2020). One specific feature bridging the effects of rapid antidepressants with sleep is the upregulation of sleep slow-wave activity (SWA), a putative marker of synaptic plasticity (Rantamäki & Kohtala, 2020). Ketamine has been shown to facilitate SWA during NREM (rapid-eye movement) sleep in rodents already in the 1990s (Feinberg & Campbell, 1995). Similar observations have been reported in a clinical study where the ketamine induced facilitation of SWA during the first NREM period was associated with clinical improvement in depressed patients (Duncan et al., 2013). We have previously proposed that rapid-acting antidepressants share the ability to induce prominent excitation and synaptic potentiation within neuronal ensembles implicated in depression (Kohtala et al., 2019; Rantamäki & Kohtala, 2020). These effects are supposedly mirrored by homeostatic emergence of SWA immediately following the treatments and again in deep sleep during which mechanisms important for sustaining the antidepressant effects are being regulated (Rantamäki & Kohtala, 2020). Since cortical excitability and synaptic strength are regulated by sleep and circadian rhythm (Kuhn et al., 2016; Ly et al., 2016; Tononi & Cirelli, 2014), the timing of treatment may contribute to the antidepressant effects. Yet, to the best of our knowledge, this issue has received minimal attention in the field. To demonstrate this discrepancy, we performed a systematic literature search on preclinical and clinical articles around rapid-acting antidepressants published during the past decade (2009-2019) and examined how they considered the circadian rhythm and sleep.

## 2. Materials and methods

The following Scopus search query was used: *“rapid-acting antidepressant*” OR “rapid acting antidepressant*” OR “fast-acting antidepressant*” OR “rapid antidepressant*” OR “electroconvulsive shock” OR “ECS” OR “ECT” OR “ketamine” OR “electroconvulsive therapy” AND “antidepressant”*. The search was further narrowed down by limiting the results to research articles published in peer-reviewed journals, English language, and publication date in range 2009-2019. Queries for preclinical (filtered results by “animal” OR “animals”) and for clinical studies (filtered results by “human” “OR “humans” OR “clinical”) were conducted on 7.8.2020, resulting in 1202 and 2379 results, respectively. The publication date range was selected based on our previous observations regarding the steep increase in ketamine-related research around 2008 (Rantamäki, 2019). With the rapid pace of increase in scientific publishing overall, more efficient and automated methods are required to curate and analyze the available knowledge (Larsen & von Ins, 2010). For example, to the purposes of this study, metadata-level information (e.g. abstract, or author-supplied and generated keywords) uncovered almost non-existent presence of sleep-related discussion in the whole dataset. On the other hand, while full-text mining of databases usually outperforms metadata-level searches, it introduces certain qualitative pitfalls by increasing the noise in the results, therefore making it more challenging to analyze and interpret (Sybrandt et al., 2018; Syed & Spruit, 2017; Westergaard et al., 2018). To overcome these limitations, a combinatory approach was utilized to limit the dataset to most relevant research of rapid-acting antidepressants using the Scopus query, and subsequently mine the full-texts for more detailed analysis of content.

The full-texts of the articles were sourced using databases available either via open access (e.g. Unpaywall; [Piwowar et al., 2018]) or University of Helsinki library. Out of the initial preclinical and clinical query results, 1168 (97 %) and 2266 (95 %), respectively, were successfully sourced and analyzed using a MATLAB script (ver. 2020b, The Mathworks Inc., Massachusetts, USA). Briefly, the documents were optimized and converted to text utilizing open source libraries (Tesseract OCR engine, Poppler) and imported to MATLAB array for cleanup of non-text characters, hyphenated words, hyperlinks, and incorrect spellings. Count of keyword “sleep” was subsequently performed using regular expression search, with validation of the result by manual count of 50 randomly selected articles. Next, we selected the 100 most cited original research papers (i.e. review papers excluded) from both preclinical and clinical domain for further analysis. Articles out of context of depression and rapid-acting antidepressants were excluded, if still present despite the initial filtering in Scopus. From clinical articles, all case reports were excluded. We chose to focus the analysis on the most cited research papers since these tend to receive greater attention among the scientific community and often guide future experimentation and build-up of scientific theories. Again, the binary inclusion of keyword “sleep” was checked from the main text, excluding references. Materials and methods, figure captions, and supplementary data were analyzed for inclusion of details related to housing light cycles, during which phase treatments were administered, and during which phase behavioral experiments were conducted. Clinical studies were also similarly reviewed for time of treatment administration.

## 3. Results

A gross keyword search for “sleep” indicated general ignorance of sleep in the studies (**Figure 1A**). Indeed, in the majority of papers the word “sleep” did not appear at all in the full text (including the references). A fraction of papers mentioned sleep 1-5 times, and considerably less 6 or more times. The same lack of acknowledgement of sleep applies also to the 100 most cited research papers **(Figure 1B)**. In fact, most of the few articles mentioning sleep do not discuss the role of sleep in the observed effects, but instead mention it in the context of treatment timing (e.g. drug being administered “after overnight sleep at clinic”) or in the introduction describing core symptoms of depression related to decline of sleep quality. Little attention was also paid to other chronobiology-related experimental details. Of the analyzed preclinical studies, only 6% reported conducting the treatments during the active period of animals **(Figure 1C)**. In 22 % of the animal studies, drug treatments were reported to be conducted during the light phase (inactive period), while most of the studies omitted the disclosure of treatment time. It can be speculated, however, that a significant proportion - if not all - of the studies omitting this information were conducted according to common practices of neuroscience during species-inappropriate time (Hawkins & Golledge, 2018). The circadian time of behavioral testing was disclosed in 36 % of the studies - with only 8 % of the experiments conducted during the active period (data not shown).

**Figure 1.**
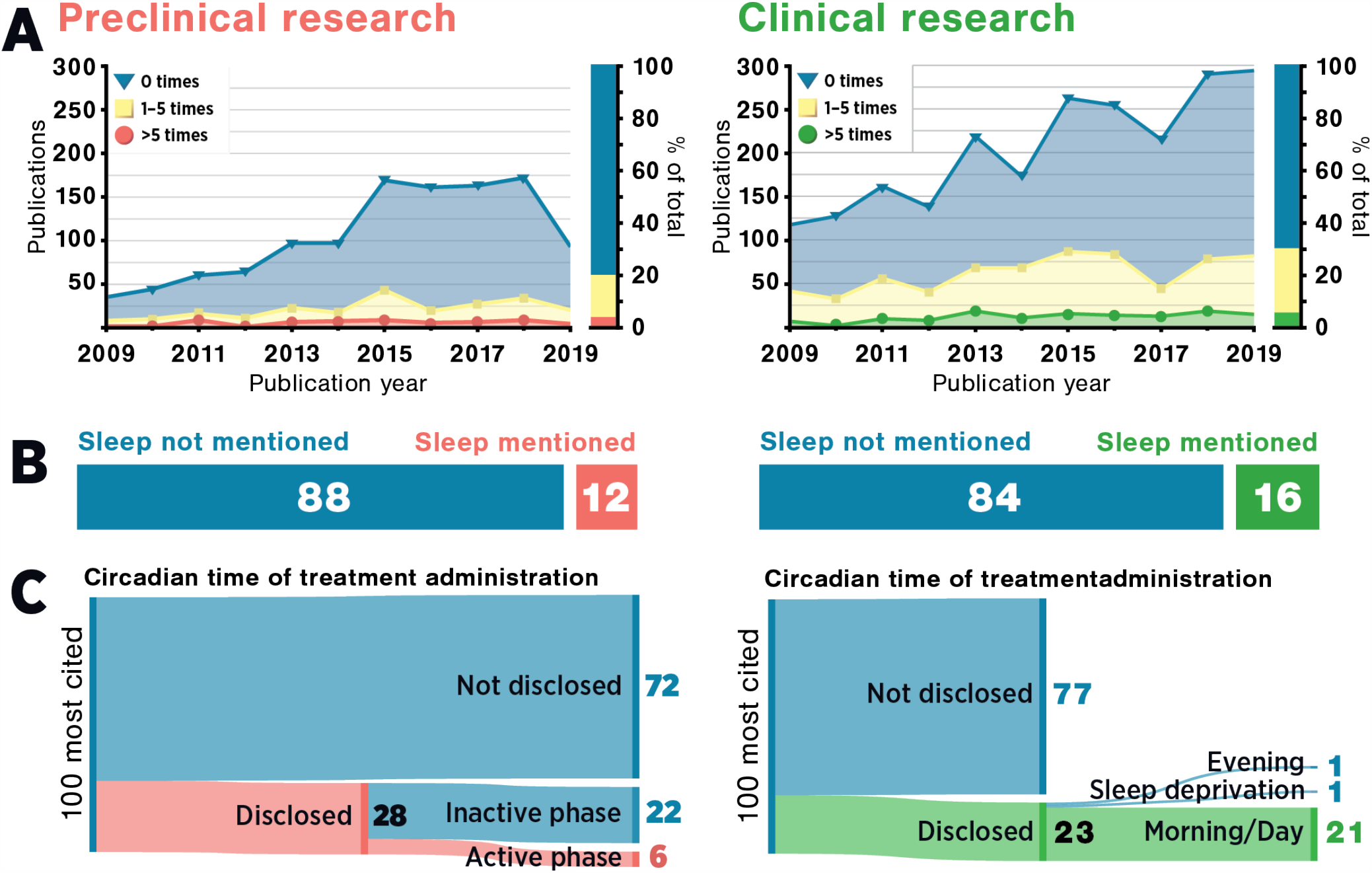
Oversight of sleep in studies of rapid-acting antidepressants published between 2009-2019. **A**) A temporal overview of the whole dataset demonstrating the count of keyword “sleep” in preclinical (n=1169; left) and clinical (n=2266; right) research papers. **B**) Proportion of top 100 cited preclinical and clinical research articles mentioning the word “sleep”. **C**) Disclosure of information regarding circadian time of treatment in the top 100 cited preclinical (left) and clinical (right) research articles.

To the best of our knowledge, no existing treatment guidelines specify the optimal time for ketamine administration. Routine practice in the clinic is, somewhat obviously, to give the treatments during “office hours”. Likely, if fasting before treatment is required (as is the case with ECT), the treatments are administered early in the day after overnight fast. Indeed, according to our clinical search results, the majority of the studies in which the treatment time was disclosed reported treating the patients either in the morning (13 %), or after overnight fast (6 %) **(Figure 1C)**. However, it should be noted that akin to preclinical studies, only 23 % of the studies disclosed the actual time of day. Three studies disclosed treating the patients in slightly differing protocols in both the morning and afternoon (Ferrucci et al., 2009), in the afternoon (Baeken et al., 2013), or surprisingly, before sleep (Irwin et al., 2013). In the last study terminally-ill patients, suffering also from depression and anxiety, received a nightly subanesthetic oral dose of ketamine, which perhaps less surprisingly led some patients reporting difficulty falling asleep. One study involved overnight sleep deprivation as the treatment, and therefore treatment time analysis is not applicable to it. The results demonstrate an evident lack of consideration for the treatment administration time in experiment planning and reporting. Most notably, the results also illustrate an alarming polarity between the administration practices of preclinical and clinical research; while patients receive the treatments during their active period, experimental animals are commonly dosed during their resting phase.

We analyzed also various other parameters present in the research articles. While the most cited preclinical research was mainly focused on ketamine, clinical research employed a larger spectrum of different treatment modalities including various brain stimulations, ketamine analogues, and combinations of methods. In preclinical studies, rats (four different strains) were the most common choice of species (data not shown). Mice, the second most employed animal, were represented by five different strains and numerous other specific genetic mutants. This might serve as a source for translational error also in the context of wake—sleep physiology, as different species and strains vary in their baseline sleep parameters, as reflected in their sensitivity to sleep deprivation (Huber et al., 2000; Wisor et al., 2008), synaptic potentiation, and antidepressant effects of ketamine (Tizabi et al., 2012). Additionally, we found that preclinical studies were characterized by primary use of male rodents. This was found to contrast the clinical studies, where female-favoring but more equal representation of both sexes was observed. Generally, the 100 most cited clinical studies were found to be small trials of less than 50 participants (data not shown).

## 4. Discussion

Sleep plays an important role in maintaining neuronal homeostasis and regulating synaptic plasticity, learning and memory. Sleep restores energy supplies, and promotes recovery from cellular stress, repair of cellular and DNA damage, clearance of metabolic byproducts, modulation of immune system, and regulation of temperature and metabolism (Bass & Takahashi, 2010; Bellesi et al., 2016; Benington & Craig Heller, 1995; Inokawa et al., 2020; Mackiewicz et al., 2007; Refinetti & Menaker, 1992; Reimund, 1994; Scharf et al., 2008; Zada et al., 2019). However, the impact of sleep or circadian biology in drug action have been only rarely considered (Ruben et al., 2019). To the best of our knowledge, the impact of circadian timing of drug administration has not been evaluated in the context of treatment effect, or potential translational bias, in studies involving rapid-acting antidepressants. In contrast, according to our focused literature review, the majority of the most cited preclinical and clinical research reports involving rapid-acting antidepressants do not consider the role of sleep (or the lack of it) in experimental design and/or obtained observations, with the majority of studies even omitting the disclosure of treatment time. Based on the results of the preclinical study search, it appears that in studies involving ketamine or other rapid-acting antidepressants involve animal handling and experimental treatments preferentially during their inactive period. This contradicts the regular sleep physiology of the animals, and is the polar opposite of clinical practice, where patients are routinely treated during “business hours” of the day. For human studies, administration during daytime aligns relatively well to the biological rhythm of the patients, and therefore any stimulatory effects from ketamine have dissipated before bedtime. On the other hand, the introduction of stimulatory treatments during subjective nighttime may disrupt physiological sleep, leading to potentially unforeseen experimental consequences. Incorrect circadian timing and circadian disruption (Karatsoreos et al., 2011) have previously been shown to induce significant variation in behavioral testing results (Cain et al., 2004; Chaudhury & Colwell, 2002; Verma et al., 2010) and effects of various pharmacological agents (Dallmann et al., 2014).

To date, there exists an ample amount of evidence linking effects of ketamine with the physiology of sleep and circadian regulation (Rantamäki & Kohtala, 2020). Circadian markers appear to have predictive value in estimation of the antidepressant and antisuicidal effects of ketamine (Duncan et al., 2017, 2018; Vande Voort et al., 2017), ECT (Kim et al., 2018; Szuba et al., 1997), and sleep deprivation (Bunney & Bunney, 2013). While there is currently no causal evidence demonstrating the convergence of these treatments on the same mechanisms in the brain, a potential shared property of rapid-acting antidepressants is their ability to transiently amplify neuronal activity in the cortex (Rantamäki & Kohtala, 2020). As evidenced by the increase in markers of synaptic plasticity and sleep SWA, increased cortical activity augments the buildup of homeostatic sleep pressure and leads to the entrainment of circadian clockwork. In this context, several recently proposed hypotheses offer noteworthy perspectives as to why sleep and circadian mechanisms should be properly accounted for in both translational animal research as well as clinical studies. For example, Wolf et al (2016) suggests that depression is characterized by deficient synaptic plasticity and the inability of synapses to reach an optimal zone for induction of LTP (long-term potentiation) during the day. Thus, prolonged waking may facilitate synaptic potentiation, subsequently resulting in a therapeutic response.

Notably, previous studies have demonstrated sleep deprivation to have surprisingly similar molecular and behavioral effects in comparison to what are commonly associated with ketamine. For example, different types of sleep deprivation have been shown to induce antidepressant-like effects lasting for days, as assayed by commonly used behavioral paradigms such as forced swimming test (Hawkins et al., 1980; Lopez-Rodriguez et al., 2004; Van Luijtelaar & Coenen, 1985). The effects of disrupted sleep are also visible in other behavioral tests related to mood and cognition (Albert et al., 1970; Hicks & Moore, 1979; Palchykova et al., 2006; Stern, 1971). Furthermore, a cornucopia of cortex-activating treatments, including various convulsant drugs (Betin et al., 1982) and optogenetic stimulation (Fuchikami et al., 2015), have been demonstrated to produce antidepressant-like and fear diminishing effects in behavioral tests, suggesting that the mechanism behind these effects at least partially involves the transient stimulation of cortical activity. In the light of these studies, it remains plausible that some effects of ketamine may be related to its ability to produce a period of “pharmacological augmentation of wake”. The excitatory and arousal-promoting effects of subanesthetic doses of ketamine have been well established (Abdallah et al., 2018; Li et al., 2016; Lu et al., 2008; Zanos et al., 2018; Zanos et al., 2018), and they lead to delayed and/or disturbed sleep when the drug is administered during the inactive period in both preclinical (Ahnaou et al., 2017) or clinical setting (Irwin et al., 2013). It is therefore conceivable that some of the effects attributed to ketamine in preclinical literature might originate from an unaccounted effect of the prolonged wake of experimental animals. This provokes the question whether the clinical treatment response would also be enhanced by delaying the administration towards the end of the subjective wake. At least, the effects of ketamine-induced arousal during the inactive period should be taken into account by, for example, including a short sleep deprivation period for all the treatment groups.

Discrepancies may also arise from the disruption of the physiological functions of sleep and the (circadian) time of administration of the interventions. These viewpoints are thoroughly discussed in the hypothesis of encoding, consolidation and renormalization in depression (ENCORE-D) (Rantamäki & Kohtala, 2020), which incorporates principal elements from the synaptic homeostasis hypothesis (SHY) of sleep (Tononi & Cirelli, 2003, 2014) and evaluates them in the context of antidepressant treatments like ketamine, ECT and sleep deprivation. One of the aspects of ENCORE-D is related to the putative renormalization of synaptic strength during sleep. Here both the accrual of synaptic potentiation during waking and the renormalization of synaptic strength during sleep offer possibilities to readjust the balance of networks that may underlie the emergence of depressive behavior. While these principles remain speculative, they offer a novel perspective to how sleep and timing may modulate antidepressant outcomes. Moreover, with recent studies demonstrating the regulation of cortical excitability by both circadian rhythmicity and sleep, it could be that treatments given in the early morning may produce different outcomes than those given late in the evening (Ly et al., 2016), further highlighting the importance of describing treatment times in both preclinical and clinical literature.

Recent research has also established a foundation for ketamine’s effects on circadian components. For example, preclinical studies have demonstrated that both sleep deprivation and ketamine influence the function of circadian molecular components, and lead to altered transcriptional clock gene output (Bellet et al., 2011; B. G. Bunney et al., 2015; Orozco-Solis et al., 2017; Wisor et al., 2008). Apart from the acute excitatory and stimulating effects of ketamine (which are powerful non-photic entraining cues for circadian rhythms), many of the molecular targets of ketamine’s acute action are also shared by the regulatory mechanisms of the mammalian circadian clock. A recent model by Duncan et al. (2017) proposes that diminished interactions between the homeostatic and circadian components of sleep promote depressed mood, and that by increasing circadian amplitude an shifting its phase, ketamine is able to restore the circadian misalignments present in depressed patients. Indeed, human studies have shown an association with ketamine’s antidepressant effects and circadian timekeeping (Duncan et al., 2017, 2018).

Besides the main focus of this review - sleep - several considerations also rise from the use of different rodent strains and from the innate differences between male and female rodents. Pharmacological studies have been traditionally conducted using male rodents, which has been suggested to produce misleading results due to prevalent sex differences (Kokras & Dalla, 2017; Raz & Miller, 2012). Sex and genetic background also lead to variability in behavioral tests (Caldarone et al., 2000; Võikar et al., 2001). Female rodents have been demonstrated to be more susceptible to the effects of ketamine in virtually all domains, including stimulant (Dossat et al., 2018), anesthetic (Winters et al., 1986), pharmacokinetic (Saland & Kabbaj, 2018), molecular (Ho et al., 2018), behavioral (Guo et al., 2016), and overall antidepressant effects (Franceschelli et al., 2015; Herzog et al., 2019; Wright & Kabbaj, 2018) (although see for example Thelen et al. (2016, 2019)). The significant sex difference in the effects of ketamine has even led some authors to call for separate reporting of data considering male and female subjects (Lees et al., 2004). Notably, clinical studies have found female brains to generally have lower excitation threshold (Sackeim et al., 1987), perhaps associated with more favorable acute treatment outcome of ECT (Clinical Psychiatry Degree, 1965; Coryell & Zimmerman, 1984; Herrington et al., 1974). The effect does not appear to be constrained to human brain, as magnetic stimulation also produces more pronounced antidepressant-like effects in female rats (Yang et al., 2007). The available literature suggests that females display overall more pronounced markers of synaptic potentiation, as evidenced by more prominent increases of SWA in response to homeostatic challenges like sleep deprivation (Armitage & Hoffmann, 2001; Manber & Armitage, 1999). The effects are unlikely restricted to ketamine only, as the sex-dependent influence of neuronal excitability appears to modulate excitatory neurotransmission (Hyer et al., 2018; Oberlander & Woolley, 2016) and plastic responses of the brain when subjected to stress and other stimuli (Armitage & Hoffmann, 2001; Farrell et al., 2015; Garrett & Wellman, 2009).

The studies briefly discussed here highlight the complexity of current research involving rapid-acting antidepressants. The literature search presented in this study demonstrates that the 200 most cited research reports in both clinical and preclinical domains - therefore perhaps the most widely read research reports of the past decade in the field of antidepressant research - display a common disregard for sleep and circadian physiology in experiment planning and reporting. We show that in preclinical research, it is customary to treat the animals using stimulatory treatments during their rest, potentially causing significant sleep disruptions. This contributes to creating a clear difference between preclinical research and standard clinical practices, but also predisposes study results to various unknown physiological and neurobiological effects, such as putative synaptic potentiation due to a form of sleep deprivation. Consideration of this is omitted - possibly due to a lack of proper understanding of the important role sleep plays in physiological processes. It is therefore conceivable that the sharp juxtaposition in the features of preclinical (experiments during inactive period, majority males), and clinical (treatments during active period, majority females) research practices could have contributed to the soaring numbers of failed clinical trials of rapid-acting antidepressants. In the light of more recent hypotheses of antidepressant actions (Duncan et al., 2019; Rantamäki & Kohtala, 2020; Zanos et al., 2018), beginning to administer the treatments during a more species-appropriate time would rather easily separate the contribution of circadian disruption from the behavioral effects observed. We wish that the data presented in this report would encourage the reader to consider the effect of sleep (or the lack of it) in their experiments or clinical practice, and to aid the research reproducibility in the field - at least by including the specific time the experiments were conducted.

## Data Availability

All the data are available upon request.

## Authorship contributions

O.A. conducted and analyzed literature search. R.S. and O.A. analyzed literature search results. O.A. prepared figures. O.A., R.S., C.Z., S.K., and T.R. wrote or contributed to the writing of the manuscript. All authors have approved the final submitted version.

